# Experimental quantification of soft tissue deformation in quasi-static single leg flexion using biplanar imaging

**DOI:** 10.1101/2021.02.15.21251415

**Authors:** Bhrigu K. Lahkar, Pierre-Yves Rohan, Jean-Jacques Yaacoub, Helene Pillet, Xavier Bonnet, Patricia Thoreux, Wafa Skalli

## Abstract

Soft tissue deformation(STD) causes the most prominent source of error in skin marker (SM) based motion analysis, commonly referred to as Soft Tissue Artifact (STA). To compensate for its effect and to accurately assess *in vivo* joint kinematics, quantification of STD in three-dimension (3D) is essential. In the literature, different invasive and radiological approaches have been employed to study how STA propagates in joint kinematics. However, there is limited reference data extensively reporting distribution of the artifact itself in 3D.

The current study was thus aimed at quantifying STD in 10 subjects along three anatomical directions. Biplanar X-ray system was used to determine true bone and SM positions while the subjects underwent quasi-static single leg flexion.

STD exhibited inter-subject similarity. A non-uniform distribution was observed at the pelvis, thigh and shank displaying maximum at the thigh (up to 18.5 mm) and minimum at the shank (up to 8 mm). STD at the pelvis and thigh displayed inter-marker similarity. STD at the pelvis was found direction independent, showing similar distribution in all the 3 directions. However, the thigh and shank exhibited higher STD in the proximal-distal direction of the bone embedded anatomical reference frame. These findings may provide more insights while interpreting motion analysis data as well to effectively strategize STA compensation methods.

## 1. Introduction

Skin Marker (SM) based motion analysis is the most common non-invasive method for estimating skeletal position and orientation in 3D space. Accuracy of such method is mainly limited by relative movement between soft tissues and the underlying bone, commonly known as Soft Tissue Artifact (STA). In order to compensate for it and to accurately estimate *in vivo* skeletal position during motion, knowledge of Soft Tissue Deformation (STD) pattern during motion is critical (Benoit et al., 2006; Stagni et al., 2005).

Several invasive (e.g., bone pins (Benoit et al., 2006; Reinschmidt et al., 1997)) and radiological studies (e.g., fluoroscopy (D’Isidoro et al., 2020; Stagni et al., 2005), biplanar X-ray (Südhoff et al., 2007; Tashman and Anderst, 2002), MRI ((Akbarshahi et al., 2010; Sangeux et al., 2006)) have been proposed to characterize STD during different motor tasks. Most of the studies concluded that STD is dependent on an individual subject, type of performed activity, marker configuration as well as locations. For instance, few studies have found that kinematic error due to STD is greater at the thigh than the shank, suggesting location- and segment-specific scheme to compensate for the artefact (Akbarshahi et al., 2010; Benoit et al., 2006; Stagni et al., 2005). Nevertheless, these studies primarily focused on quantifying the kinematic errors caused by STD rather than STD itself.

As far as the authors are aware of, one study dealt with quantification of STD at different marker locations and directions in 20 healthy volunteers (Gao and Zheng, 2008). But, due to technical limitations preventing access to the bone position, STA quantification was reported as inter-marker movement instead of marker movement relative to true bone positions. Hence, there is still a lack of reference data on subject-, location- and direction-specific STD, which may provide insight for effective STA compensation strategies for SM based motion analysis.

Amongst the different methods devised for compensating STA, multi-body optimization (MBO) method is increasingly used. It generally assigns a weight matrix reflecting the STA error distribution among the markers adhered to a segment (Lu and O’Connor, 1999). Moreover, recently our group has proposed a finite element (FE) based novel approach to compensate for STA of the lower limb and successfully evaluated in a population of 66 subjects (Lahkar et al., 2020, under review). The FE model facilitates to incorporate STA correction stiffness at each marker location, and stiffness can be calibrated based on information of local STD at each marker location and along each anatomical direction. However, owing to lack of STD data, arbitrary values were assigned for the stiffness parameters.

The current study was thus aimed at quantifying soft tissue deformation on the pelvis, thigh and shank at each marker location and in three anatomical directions during single-leg quasi-static knee flexion using low dose biplanar radiography.

## 2. Materials and methods

### 2.1 Data collection

The retrospective data included in the study recruited ten volunteers (age range: 23-40 years; weight range: 63-89 kg, height range: 1.7-1.9 m), 6 months after ACL reconstruction following approval of a relevant ethical committee. Patients with a large osteochondral defect (>1cm^2^), operated for a meniscal suture and multi-ligament knee injury, or diagnosed with a neuromuscular disorder which could impair motion, were excluded from this study. The mean IKDC (International Knee Documentation Committee) score for the subjects was 79.7±7.2. This score ranges from 0 to 100, with higher scores representing lower levels of symptoms and higher levels of function and sports activity (Irrgang et al., 2001).

Subjects were equipped with a total of 20 retro-reflective skin markers (pelvis: 4, thigh: 8 and shank: 8) according to the Plug-in Gait^®^ method (Davis et al., 1991). Three pairs of bi-planar radiographs (EOS Imaging, France) were acquired in three configurations for each subject (Fig. 1). First, a pair of radiograph was taken in the free-standing position. Then, two sequential pairs of radiographs at approximately 20° and 40° of knee flexion were acquired while each subject performed a quasi-static single-leg knee flexion. For the sake of clarity, three sequential postures will be hereafter termed as respectively pose 1, pose 2 and pose 3 for free-standing, 20° and 40° of knee flexion.

**Fig. 1.**
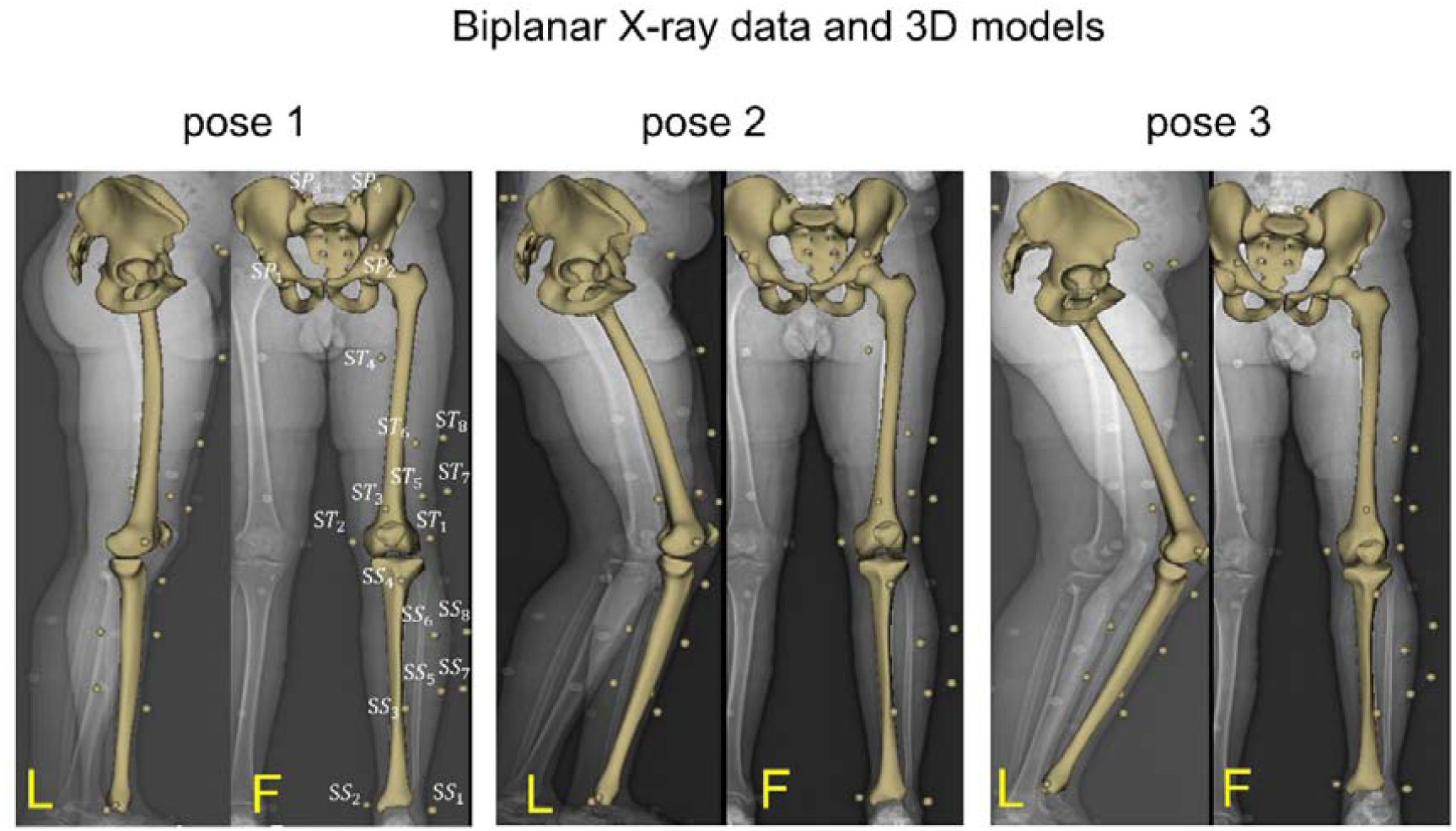
3D digital models of the pelvis, femur and tibia and their respective skin adhered markers at positions: pose 1 (free standing), pose 2 (20° knee flexion) and pose 3 (40° knee flexion) built from orthogonal radiographs. Marker nomenclature is shown at pose 1 for the pelvis: *SP*_1_ to *SP*_4_, for the thigh *ST*_1_ to *ST*_8_ and for the shank: *SS*_1_ and *SS*_8_. L: lateral and F: frontal view

3D digital models of bones (pelvis, femur and tibia) were first obtained at free-standing position using a 3D reconstruction algorithm developed previously by (Chaibi et al., 2012) for femur and tibia and (Mitton et al., 2006) for the pelvis. The 3D models were then projected on the frontal and lateral radiographs. The positions of the bony contours were manually adjusted until the contours exactly matched those of the radiographs at each pose. 3D location of skin markers at each pose was also computed from biplanar radiographs using the same procedure. Anatomical reference (R_*anat*_) frames for the femur and tibia was defined following the definition reported in (Schlatterer et al., 2009), and for the pelvis, in (Dubois, 2014). *x, y* and *z* axes of the R_*anat*_ frames are along antero-posterior, proximal-distal and medial-lateral direction respectively.

### 2.2 Quantification of STD

STD quantification on the pelvis, thigh and shank was performed based on two different schemes in line with the literature.

First, as a Soft Tissue Element (STE) deformation at each marker location as introduced in our previous work (Lahkar et al., 2020, under review). The overall procedure is briefly explained and illustrated in figure 1 below.

3D position of the skin markers (S_*i*_) in all the poses were first computed from the biplanar X-ray data and expressed in the respective bone R_*anat*_ frames. From the skin markers, a set of virtual markers referred to as subcutaneous markers (SC_*i*_), were defined 1 mm beneath the skin marker following the methodology elaborated in (Lahkar et al., 2020, under review) and illustrated in Fig. 2(A). All the soft tissue deformation effect at the marker level is reported to the STE, which connects the *S*_*i*_ to the corresponding SC_*i*_. The connection between the SC_*i*_ and the corresponding bone segment was assumed to be rigid. Due to the rigidity assumption, the locations of the subcutaneous markers in R_*anat*_ frame remained the same in all the poses (SC_1_=SC_2_ =SC_3_). Thus the absolute differences between skin and subcutaneous marker locations at pose 2 and pose 3 were computed and expressed in R_*anat*_ frames along *x, y* and *z* direction (Fig. 2(B) and 2(C)). Eventually, from the directional components, Euclidean distances were computed using equation 1.

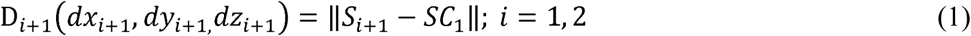

**Fig. 2.**
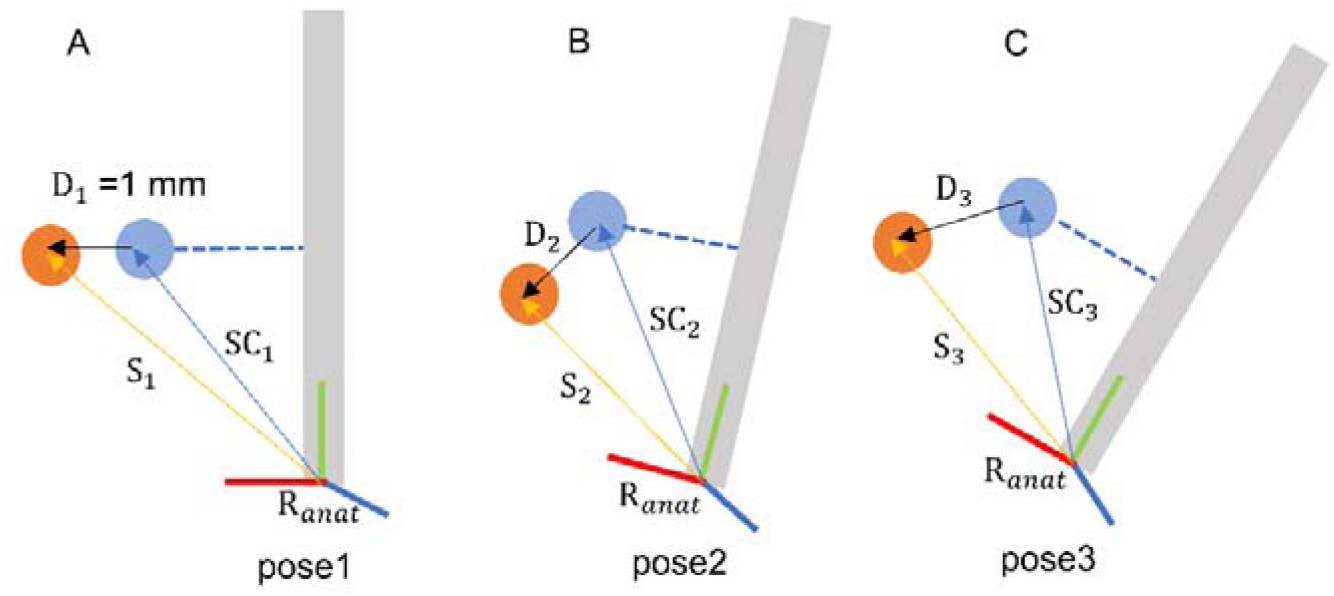
Scheme 1: Schematic representation of Soft Tissue Element (STE) deformation. Skin {S_*i*_ (*i* = 1,2,3)} and subcutaneous marker (SC_*i*_) locations are expressed in bone R_*anat*_ frames in all the poses. D_1_(1 mm), D_2_ and D_3_ are Euclidean distances between skin and subcutaneous marker at pose 1, pose 2 and pose 3 respectively. Shown only for a single marker.

Second, STD was computed as a relative displacement of the skin markers at pose 2 and pose 3 with respect to pose 1 (reference pose) as illustrated in figure 3. From the directional components, Euclidean distances were computed using equation 2.

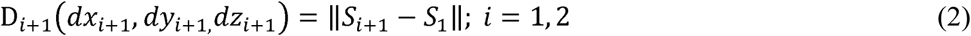

where S_*i*_ is the 3D location of the skin marker at pose *i* obtained from biplanar X-ray data and expressed in bone R_*anat*_ frames. D_*i*+1_ is the relative displacement of the skin markers at pose (i+1).

**Fig. 3.**
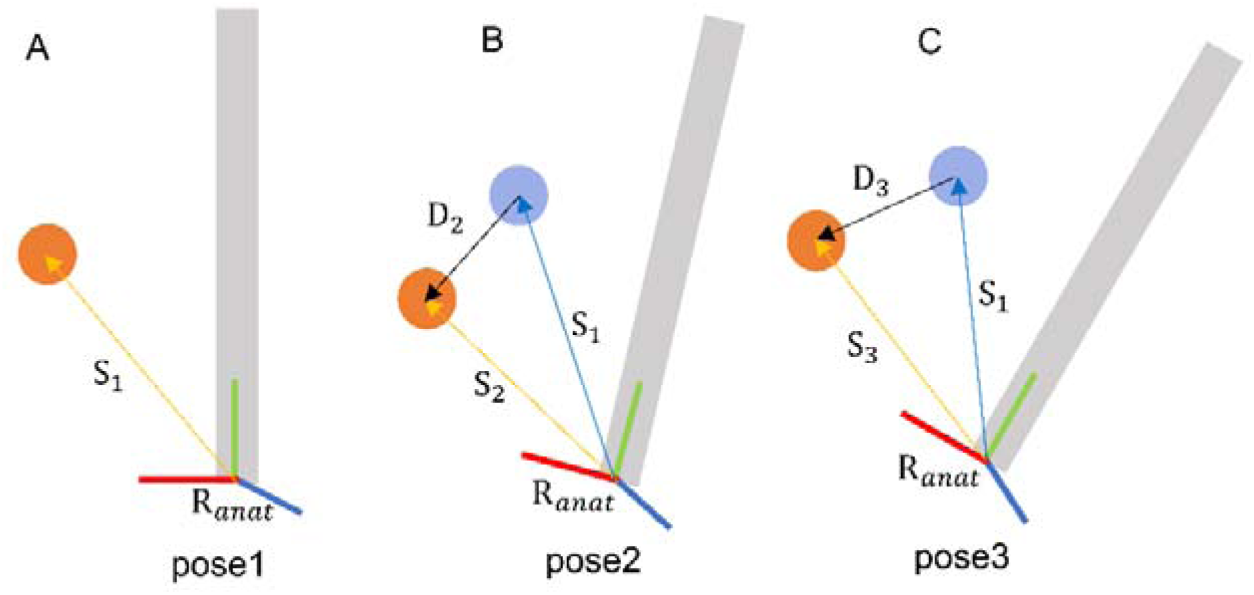
Scheme 2: STA as a skin marker relative displacement. Schematic representation of skin marker {S_*i*_ (*i* = 1,2,3)} locations expressed in bone R_*anat*_ frames in all the poses. Shown only for a single marker.

STA quantification in both the schemes was performed using a customized Matlab routine (Mathworks, Massachusetts, United States).

### 2.3 Statistical analysis

Statistical analysis on the collected data was performed using both the schemes to test 4 hypotheses.

#### STD is subject-specific

Deformation data at all marker locations were pooled together **per subject** per pose to check inter-individual similarity/variability.

#### STD is segment-specific

Deformation data for all the subjects and at all marker locations **per segment** per pose were pooled together to check inter-segment variability/similarity among pelvis, thigh and shank.

#### STD is location-specific

Deformation data for all the subjects **per marker location** per pose within a segment were pooled together to check inter-maker location variability/similarity within segments.

#### STD is direction-specific

Deformation component **in a particular anatomical direction** (*x, y* or *z*) per pose, for all the subjects and at all marker locations within a segment were pooled together to check if deformation is dependent on anatomical directions within segments.

Normality of the distributions were first assessed using the Shapiro-Wilk test. According to the outcomes of normality test, ANOVA or nonparametric Kruskal-Wallis (KW) test was performed to observe intergroup differences using the built-in MATLAB functions. We also performed pairwise comparisons with Student t-test or nonparametric Man-Whitney *U* test (with Bonferroni’s correction). For all the tests, the significance level was set to 0.05 (*) and 0.01 (**) *a priori*.

## 3. Results

Results obtained with both the schemes were similar, with mean differences between the schemes less than 1 mm (appendix 1). Therefore, the results of the statistical analysis are shown only for scheme 1, where deformation at each marker location is presented as STE deformation.

Figure 4 illustrates STD (i.e., median, quartiles, minimum, maximum and outliers) per subject (*P*_1_ to *P*_10_) per pose. Results of the KW test showed, the null hypothesis that STD for each subject comes from the same distribution cannot be accepted (*p*<0.05). The pairwise test showed that there is a significant difference in STD between subjects *P*_2_ and *P*_5_ at pose 2. Similarly, subjects *P*_1_ and *P*_6_, and *P*_4_ and *P*_6_ displayed significantly dissimilar STD at pose 3 only. Subjects *P*_1_-*P*_4_ and *P*_7_-*P*_10_ showed inter-subject similarity among them. Overall, higher STD was observed for the subjects *P*_2_, *P*_6_ and *P*_7_ at pose 3, exhibiting maximum value up to 45 mm for *P*_7_.

**Fig. 4.**
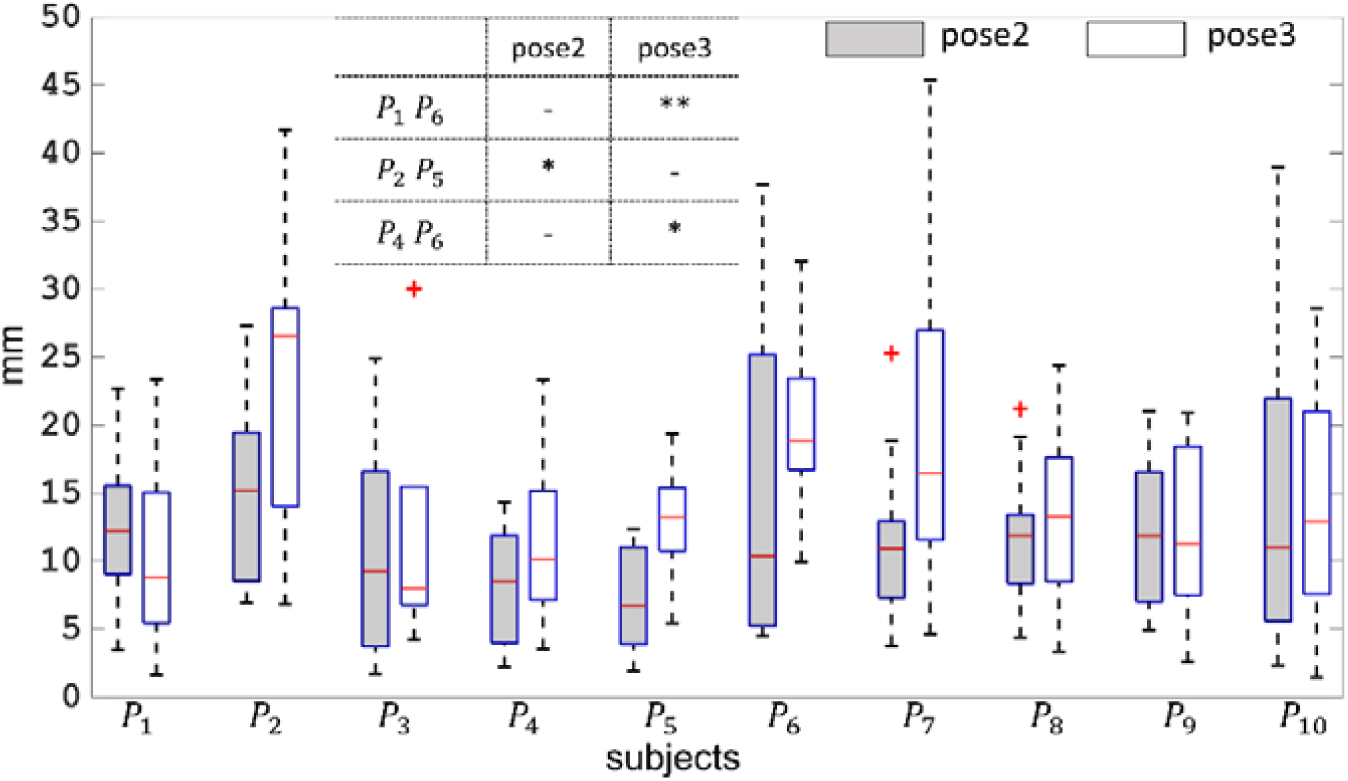
Boxplot for subject-specific STD presented at pose 2 and pose 3. *P*_1_ to *P*_10_ are number of subjects. Only significant parameters are presented in the table.

Figure 5 represents STD per segment per pose for all the subjects. KW test revealed that STD across all the segments was distinctly different (*p*<0.05). Among the segments, STD for the thigh was observed significantly higher at both the poses with values (median) 13.5 mm and 18.5 respectively. Lowest STD (median: 5 mm) was observed for the shank at pose 2. STD at the pelvis was found around 13 mm (median). Few outliers were observed at both the poses, particularly for the thigh and shank.

**Fig. 5.**
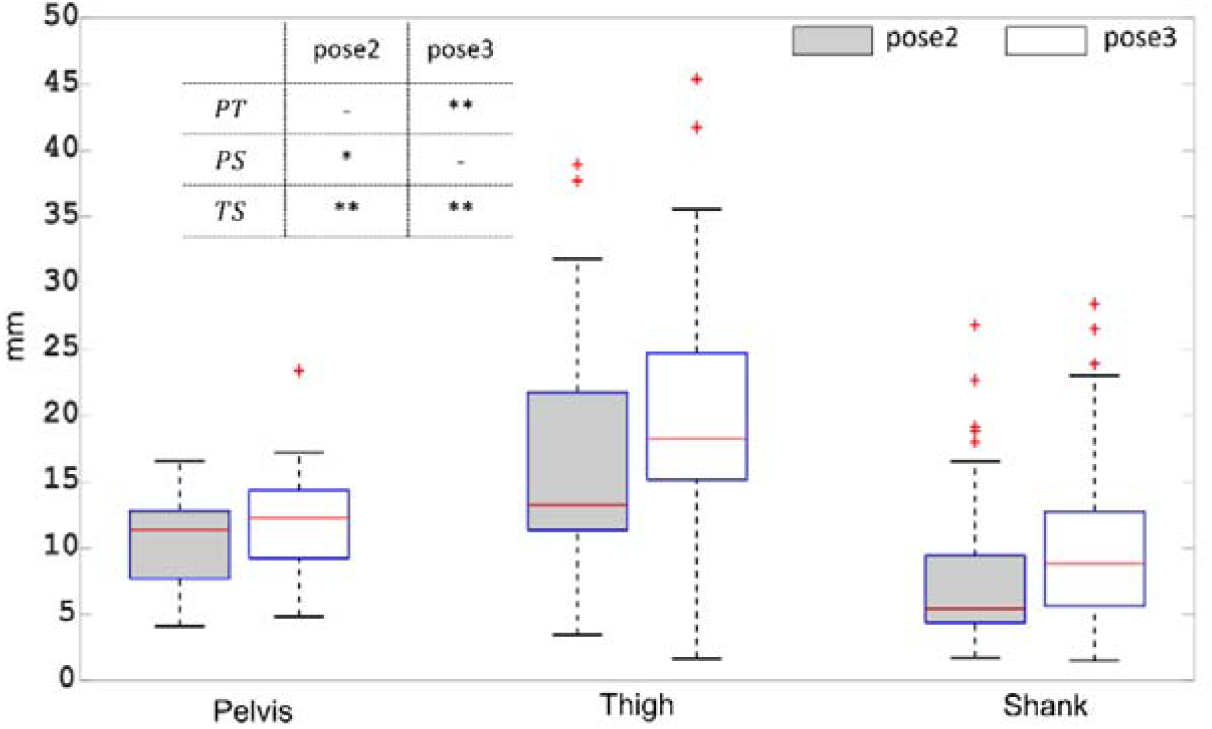
Boxplot for segment-specific STA presented at pose 2 and pose 3. Only significant parameters are presented in the table. P: Pelvis, T: Thigh and S: Shank.

Figure 6 depicts STD at each marker location (left column figures) and per anatomical direction (right column figures) within a segment. In the case of location-specific analysis, STD at each marker location of pelvis appeared similar (*p*>0.05) for both the poses with values (median) within 11 mm to 13 mm. Similarly, for the shank, no significant difference in STD among different locations were seen (*p*>0.05). For the thigh, only *ST*_1_ and *ST*_4_ displayed significantly dissimilar STD at both the poses, with the highest STD (median: 26 mm) for *ST*_4_ and the lowest (median: 15 mm) for *ST*_1_ at pose 3.

**Fig. 6.**
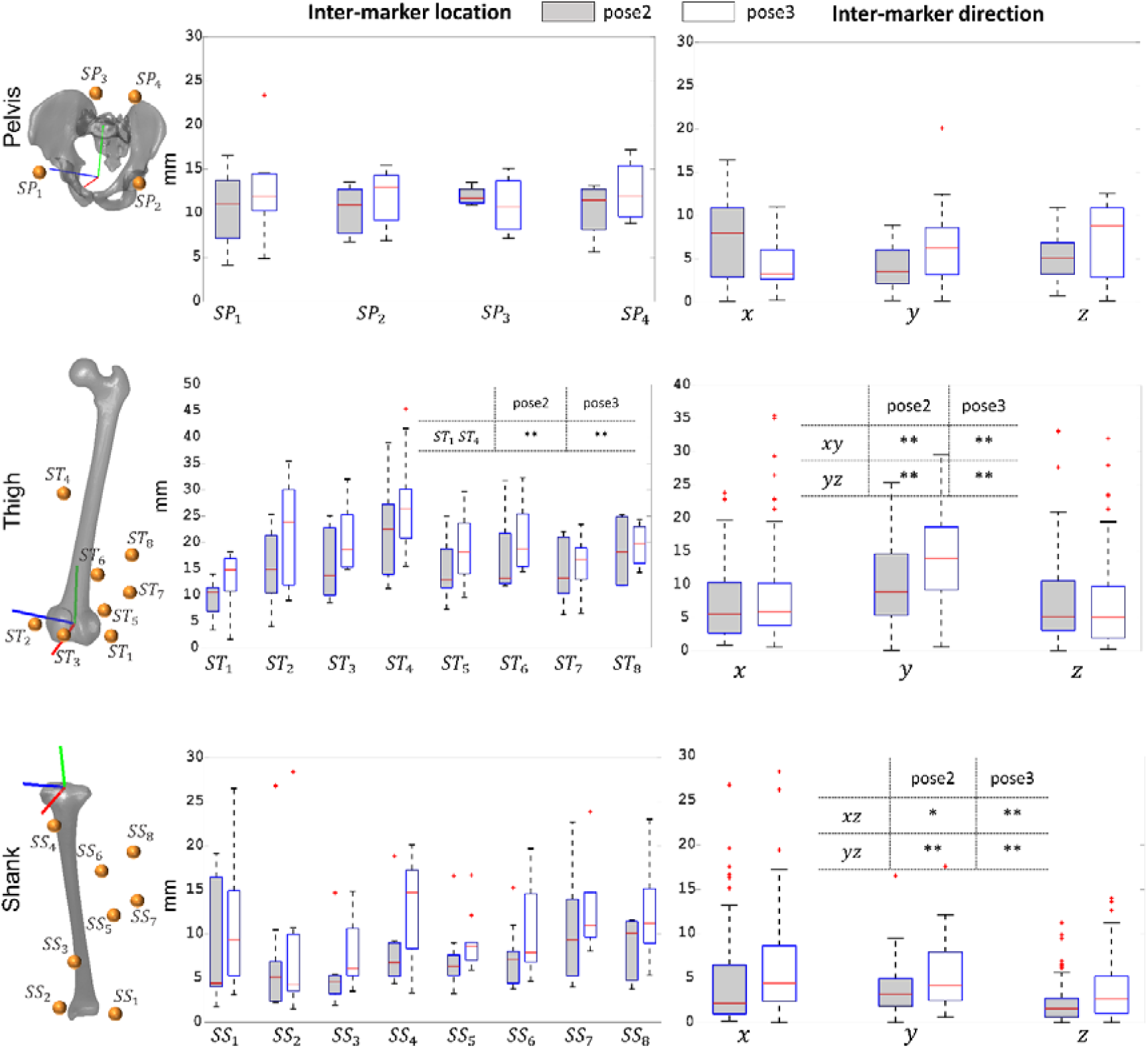
Boxplot for location-specific STD (left column) and direction-specific (right column) presented at pose 2 and pose 3 for the pelvis, thigh and shank. Only significant parameters are presented in the tables. Anatomical reference frames (R_*anat*_) are also highlighted on the bone segments in different colours according to different axes. Green: proximal-distal direction, red: anter-posterior direction, blue: lateral-medial direction.

In the case of direction-specific analysis, a similar (*p*>0.05) STD was observed across the three anatomical directions of the pelvis R_*anat*_ frame. STD along proximal-distal direction (in bone embedded R_*anat*_ frame) of the thigh was observed distinctly higher in both the poses, while showing similar values along antero-posterior and lateral-medial direction. STD at the shank was appeared similar along antero-posetrior and proximal-distal direction, while revealing significantly lower values along the medial-lateral direction. Overall analysis showed higher STD at pose 3 as compared to pose 2.

## 4. Discussion

In order to compensate for STA and to interpret SM-based motion analysis data, knowledge of Soft Tissue Deformation (STD) pattern in 3D is essential. Yet, there is a paucity of reference data in the literature comprehensively showing variability of STD among individuals, segments, marker locations and along the three anatomical directions. The purpose of this study was to quantify STD in 3D at the pelvis, thigh and shank for 10 subjects. Two schemes were employed to quantity STD, although exhibited similar results.

The rationale behind employing two schemes is that both of them may serve two different communities; first, those particularly deal with compensation methods, and second those deal with STD quantification. The first scheme intends to address conventional STA compensation methods (such as MBO) that generally minimizes measured and model determined marker position. The subcutaneous markers are analogous to the model determined markers. This approach could also be helpful for finite element-based STA compensation method, where a deformable element connecting the skin and subcutaneous marker accounts for all soft tissue deformation (Lahkar et al., 2020, unpublished). The second scheme pertains to STD quantification methods, where soft tissue deformation is considered as a relative displacement of skin markers at different poses with respect to the reference pose.

Overall, soft tissue deformation displayed inter-subject similarity in most of the subjects showing a similar STD pattern. Although such observation is in contrast to the current prevailing idea of STD as subject-specific, yet found in accordance with one study which explained overshadowing of similarity by dissimilarity for few subjects (Gao and Zheng, 2008). Secondly, segment-specific STD was observed exhibiting the highest deformation at the thigh followed by the pelvis and the shank (Akbarshahi et al., 2010; Walker, 2015). A similar observation was also reported in studies that measured higher kinematic error at the thigh (Sangeux et al., 2006; Stagni et al., 2005).

STD at the pelvis and shank exhibited no inter-marker variability. For the thigh, the marker (*ST*_4_) placed towards the hip joint showed significantly higher STD, where muscle thickness is higher (Rouhandeh and Joslin, 2018). Except *ST*_4_, other markers at the displayed similar STD.

STA occurred in all the three directions of the bone embedded anatomical frames, however not uniform for the thigh and shank in particular. Soft tissue deformation in proximal-distal direction of the thigh and shank was distinctly higher. This is probably due to the orientation of the muscular structure of the thigh and shank, which contracts and relaxes during movement along its length. Deformation in the medial-lateral direction was noticed the lowest. A similar observation was also reported in the literature (Gao and Zheng, 2008).

This study, to the authors’ knowledge, is the second attempt to use EOS low dose system, allowing to quantify STD in 3D. Previously, our group used EOS to investigate motion of lower limb attachment systems with respect to the underlying bone (Südhoff et al., 2007). It is to be noted that because of the limited acquisition volume within the EOS, markers present in the radiographs were not consistent throughout the subjects. Few markers couldn’t be located in the radiographs of some subjects either in the orthogonal views or in the consecutive poses. Moreover, both due to limited acquisition volume and ethical reasons, the number of poses had to be limited. Also, we acknowledge that STD reported in this study doesn’t include inertial effects, as the movement under consideration was quasi-static. Quantified STD is a consequence of both muscle contraction and skin sliding. Currently, other existing methods, such as invasive attachments and fluoroscopic measurements, have been shown useful to quantify soft tissue deformation. But, invasive methods are prone to alter free soft tissue movement and therefore, may impact its results. Fluoroscopy is also not effective for capturing the entire lower limb, although efficient for local observations in dynamics. Hence, EOS in conjunction with skin markers can serve as a gold standard to locate actual bone positions as well to quantify STD for a limited range of motion.

The findings in the study may open up effective STA compensation strategies for SM-based motion analysis. Instead of assigning arbitrary STA correction stiffness, subjects with similar STD patterns can be grouped together to assign the same correction stiffness. Moreover, for such quasi-static activities, all the markers at the pelvis can be grouped together to assign the same stiffness values. Similar is the case for the shank. For the thigh, marker locations where STD was observed highest can be assigned with the lowest stiffness and vice versa. To be noted that while assigning stiffness values for the thigh and shank, different stiffness values need to be defined along different anatomical direction. In conclusion, although the STD data provided in this study may be beneficial for future STA compensation approaches, further study would be required in different dynamic activities.

## Data Availability

Data can be provided subject to request

## Acknowledgments

The authors are deeply grateful to the ParisTech BiomecAM chair program on subject-specific musculoskeletal modeling for financial support.

## Appendix 1

**Scheme 1.**
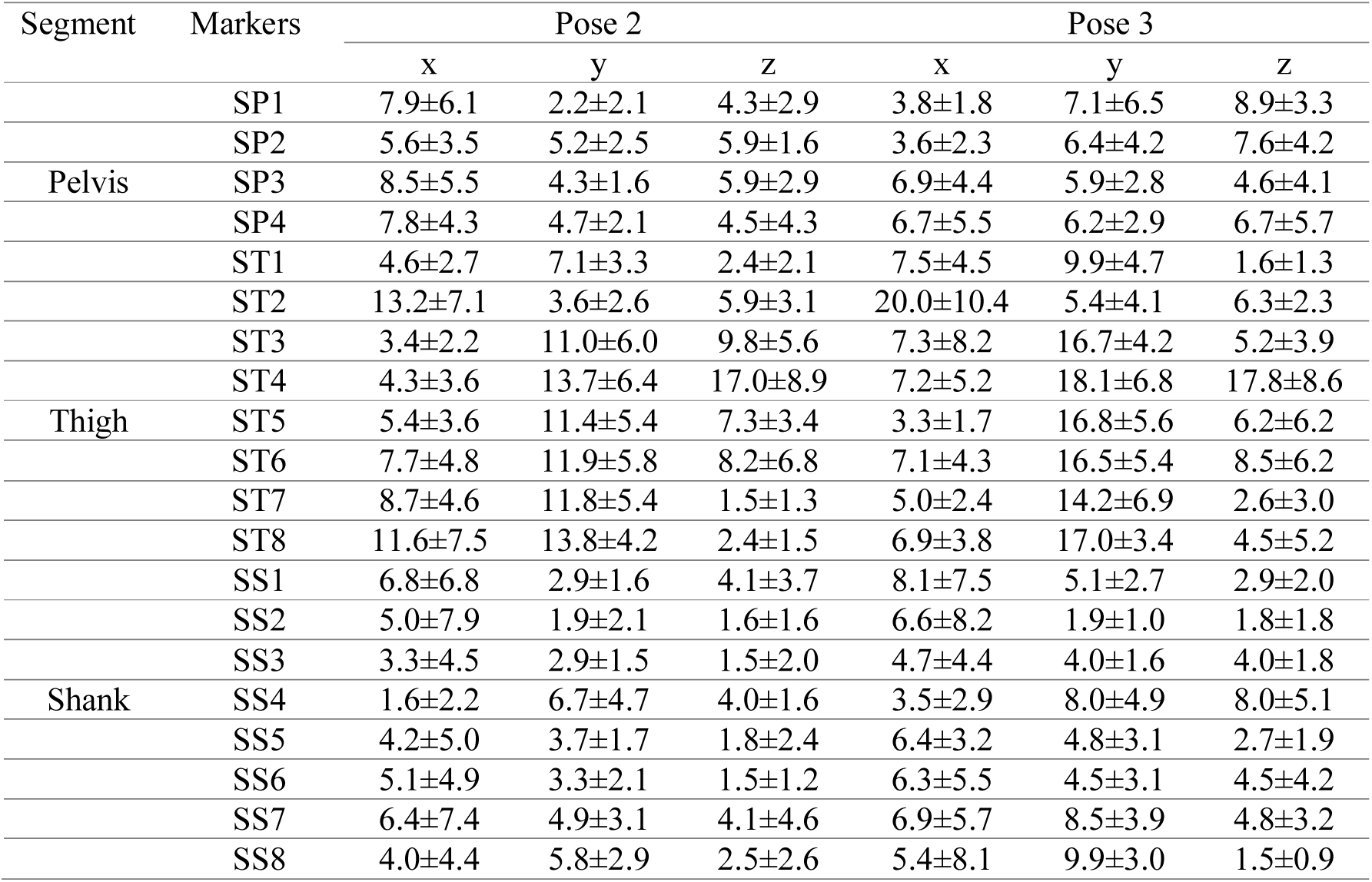
STD at pose 2 and pose 3 presented as Mean±1SD at each marker location and along x,y and z direction of R_*anat*_ frame

**Scheme 2.**
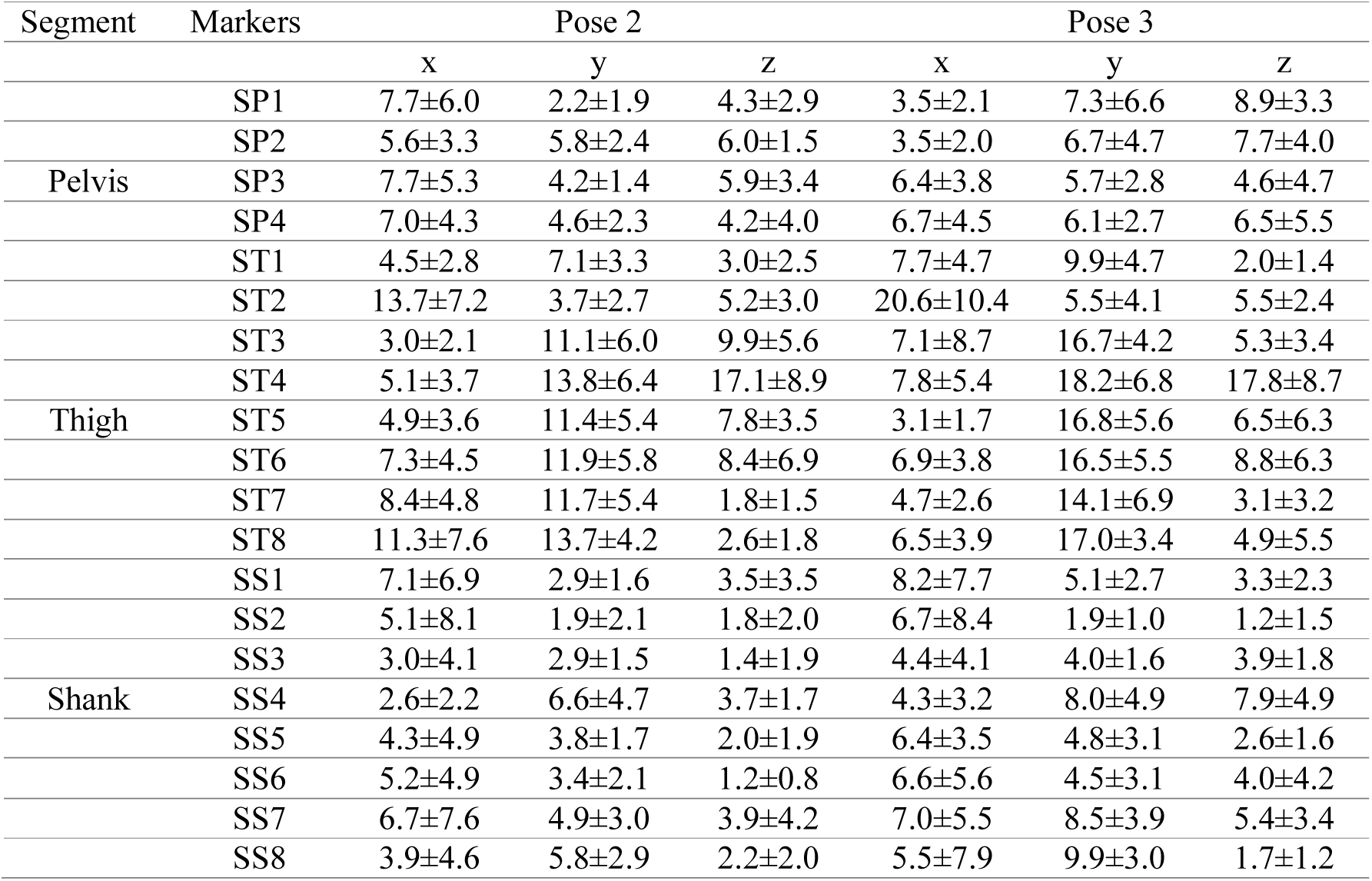
STD at pose 2 and pose 3 presented as Mean±1SD at each marker location and along x,y and z direction of R_*anat*_ frame

## References

Akbarshahi, M., Schache, A.G., Fernandez, J.W., Baker, R., Banks, S., Pandy, M.G., 2010. Non-invasive assessment of soft-tissue artifact and its effect on knee joint kinematics during functional activity. J Biomech 43, 1292–1301. https://doi.org/10.1016/j.jbiomech.2010.01.002

Benoit, D.L., Ramsey, D.K., Lamontagne, M., Xu, L., Wretenberg, P., Renström, P., 2006. Effect of skin movement artifact on knee kinematics during gait and cutting motions measured in vivo. Gait Posture 24, 152–164. https://doi.org/10.1016/j.gaitpost.2005.04.012

Cappozzo, A., Catani, F., Leardini, A., Benedetti, M.G., Della Croce, U., 1996. Position and orientation in space of bones during movement: Experimental artefacts. Clin Biomech 11, 90– 100. https://doi.org/10.1016/0268-0033(95)00046-1

Chaibi, Y., Cresson, T., Aubert, B., Hausselle, J., Neyret, P., Hauger, O., de Guise, J.A., Skalli, W., 2012. Fast 3D reconstruction of the lower limb using a parametric model and statistical inferences and clinical measurements calculation from biplanar X-rays. Comput Methods Biomech Biomed Engin 15, 457–466. https://doi.org/10.1080/10255842.2010.540758

D’Isidoro, F., Brockmann, C., Ferguson, S.J., 2020. Effects of the soft tissue artefact on the hip joint kinematics during unrestricted activities of daily living. J Biomech 109717. https://doi.org/10.1016/j.jbiomech.2020.109717

Davis, R.B., Õunpuu, S., Tyburski, D., 1991. A gait analysis data collection and reduction technique. Hum Mov Sci 10, 575–587. https://doi.org/10.1016/0167-9457(91)90046-Z

Dubois, G., 2014. Contribution à la modélisation musculo-squelettique personnalisée du membre inférieur combinant stéréoradiographie et ultrason.

Gao, B., Zheng, N. (Nigel), 2008. Investigation of soft tissue movement during level walking: Translations and rotations of skin markers. J Biomech 41, 3189–3195. https://doi.org/10.1016/j.jbiomech.2008.08.028

Irrgang, J.J., Anderson, A.F., Boland, A.L., Harner, C.D., Kurosaka, M., Neyret, P., Richmond, J.C., Shelborne, K.D., 2001. Development and validation of the international knee documentation committee subjective knee form. Am J Sports Med 29, 600–613. https://doi.org/10.1177/03635465010290051301

Lu, T.W., O’Connor, J.J., 1999. Bone position estimation from skin marker co-ordinates using global optimisation with joint constraints. J Biomech 32, 129–134. https://doi.org/10.1016/S0021-9290(98)00158-4

Mitton, D., Deschênes, S., Laporte, S., Godbout, B., Bertrand, S., de Guise, J.A., Skalli, W., 2006. 3D reconstruction of the pelvis from bi-planar radiography. Comput Methods Biomech Biomed Engin 9, 1–5. https://doi.org/10.1080/10255840500521786

Reinschmidt, C., Van Den Bogert, A.J., Nigg, B.M., Lundberg, A., Murphy, N., 1997. Effect of skin movement on the analysis of skeletal knee joint motion during running. J Biomech 30, 729–732. https://doi.org/10.1016/S0021-9290(97)00001-8

Rouhandeh, A., Joslin, C., 2018. Soft-tissue artefact assessment and compensation in motion analysis by combining motion capture data and ultrasound depth measurements. VISIGRAPP 2018 - Proc 13th Int Jt Conf Comput Vision, Imaging Comput Graph Theory Appl 4, 511–521. https://doi.org/10.5220/0006624205110521

Sangeux, M., Marin, F., Charleux, F., Dürselen, L., Ho Ba Tho, M.C., 2006. Quantification of the 3D relative movement of external marker sets vs. bones based on magnetic resonance imaging. Clin Biomech 21, 984–991. https://doi.org/10.1016/j.clinbiomech.2006.05.006

Schlatterer, B., Suedhoff, I., Bonnet, X., Catonne, Y., Maestro, M., Skalli, W., 2009. Skeletal landmarks for TKR implantations: Evaluation of their accuracy using EOS imaging acquisition system. Orthop Traumatol Surg Res 95, 2–11. https://doi.org/10.1016/j.otsr.2008.05.001

Stagni, R., Fantozzi, S., Cappello, A., Leardini, A., 2005. Quantification of soft tissue artefact in motion analysis by combining 3D fluoroscopy and stereophotogrammetry: A study on two subjects. Clin Biomech 20, 320–329. https://doi.org/10.1016/j.clinbiomech.2004.11.012

Südhoff, I., Van Driessche, S., Laporte, S., de Guise, J.A., Skalli, W., 2007. Comparing three attachment systems used to determine knee kinematics during gait. Gait Posture 25, 533–543. https://doi.org/10.1016/j.gaitpost.2006.06.002

Tashman, S., Anderst, W., 2002. Skin motion artifacts at the knee during impact movements. Proc 7th Annu Meet Gait Clin Mov Anal Soc.

Walker, P.S., 2015. The design and pre-clinical evaluation of knee replacements for osteoarthritis. J Biomech 48, 742–749. https://doi.org/10.1016/j.jbiomech.2014.12.012

